# Transdiagnostic similarities and distinctions in brain networks associated with ASD symptoms: A prospective cohort study

**DOI:** 10.1101/2025.09.17.25336005

**Authors:** Jennifer L. Bruno, Julia R. Plank, Sam Leder, Evelyn MR Lake, Emily S. Finn, Tamar Green

**Author notes:** Corresponding author. 1520 Page Mill Road Palo Alto, CA 94304. Authors contributed equally (Jennifer L. Bruno, Julia R. Plank).

## Abstract

**Background:** Despite high rates of autism spectrum disorder (ASD), understanding of pathophysiology is limited. The RAS-mitogen-activated protein kinase (RAS-MAPK) pathway plays a crucial role in ASD and is altered in children with Noonan syndrome (NS). Children with NS offer a unique model to disentangle genetic and neurological underpinnings of ASD.

**Methods:** This study aimed to examine functional brain network anatomy underlying ASD symptoms in children with NS (n=28, mean age=8.24), and tested generalizability of models developed in a non-syndromic cohort enriched for ASD (Autism Brain Imaging Data Exchange (ABIDE), n=352, mean age=11.0). Connectome-based predictive modeling (CPM) was applied to fMRI data to predict the severity of autism symptoms, indexed by the Social Responsive Scale (SRS), in children with NS. Next, we tested if a model developed to predict autism symptoms in an autism-enriched sample of children without genetic diagnosis (ABIDE) could predict autism symptoms in children with NS.

**Results:** Predicted SRS scores were significantly associated with observed SRS scores in NS (*r_s_*=0.43, *p*=.011). Application of the predictive model generated in the autism-enriched cohort (ABIDE) significantly predicted observed SRS scores in NS (*r_s_*=0.460, *p*=.018). Predictive brain networks in both NS and the non-syndromic cohorts included subcortical-cerebellar networks and visual processing networks.

**Limitations:** The size of our NS cohort is small, given the rarity of NS. However, the significant cross-dataset comparison yielded in this study suggests that use of large publicly available datasets can be useful in contextualizing smaller and harder to collect datasets in rare genetic syndromes.

**Conclusions:** The presence of shared brain networks suggests a converging pattern of functional connectivity underlying autism symptoms, irrespective of genetic diagnosis. Evidence of shared brain networks in children with idiopathic autism and NS highlights the role of RAS-MAPK in autism symptoms and points to the value of leveraging human genetic models to enhance our understanding of idiopathic ASD.

## Background

Autism Spectrum Disorder (ASD) is a complex neurodevelopmental disorder characterized by deficits in social communication and restricted and repetitive behaviors (1). One in 36 children U.S. meet criteria for ASD diagnosis (2), and increasing prevalence has been observed worldwide (3). Despite ongoing research efforts to identify the brain basis of ASD (4), clinical care for children with ASD are limited due to challenges with accurate diagnosis, early identification and prediction of outcomes. This is partly due to the heterogeneity of ASD (5) and to the limitations of neuroscience methods used to date.

Understanding the genetic underpinnings of ASD holds potential to resolve challenges with heterogeneity and make strides toward understanding underlying pathophysiology. ASD is highly heritable and, most often, the genetic risk is conferred by common variants each with a small effect that cumulatively increase one’s risk (6). While the genes implicated in ASD risk are diverse, they cluster in six main functional pathways (7,8). One of these pathways is the RAS-mitogen-activated protein kinase (RAS-MAPK) pathway which was originally identified for its role in oncogenesis (9). The RAS-MAPK pathway has since been increasingly recognized for its key role in brain development and in deviation from typical development (10,11). Examining children with RASopathies, genetic disorders associated with alterations along the RAS-MAPK pathway (12), is a translatable strategy for understanding the role of this pathway in ASD, and, when compared and contrasted with idiopathic (non-syndromic) ASD, offers a unique lens with which to disentangle complex genetic and neurological underpinnings of ASD symptoms.

Noonan syndrome (NS) occurs in 1:2000 and is the most common RASopathy. NS is caused most often by a germline mutation in the *PTPN11* or *SOS1* gene (∼60% of cases) which in turn upregulates the RAS-MAPK signaling cascade (13,14). Children with NS consistently exhibit social deficits (15–17), such as difficulty maintaining friendships as well as rigid and repetitive behaviors, sensory sensitivity, and restricted interests (14). Twelve to thirty percent of children with NS meet diagnostic criteria for ASD (14,18,19). Existing neuroimaging literature indicates overlap across structural (20–22) and functional (23–25) brain mechanisms in NS and idiopathic ASD (iASD). However, studies in genetic model syndromes such as NS have limited sample sizes, due to the rare occurrence of the syndromes, thus computational modeling approaches suffer from overfitting and results lack generalizability.

The present study seeks to improve the clinical translational utility of neuroscience research by combining a genetics-first approach, in which we examine individuals with NS as a model for ASD, with large scale data and computational modeling to improve generalizability. The overarching goal is to identify functional brain networks that underlie ASD symptoms in children with and without NS. Identifying similarities and differences will provide clues as to how much of the underlying neural endophenotypes are similar across children with NS, children with non-genetic iASD and in children who are typically developing (TD). Linking neural circuitry with underlying ASD symptoms may help inform the development of personalized treatment approaches. Including individuals with neurogenetic syndromes such as NS in mechanistic research, such as the present study, is also imperative to facilitate their inclusion and representation in future work.

We applied connectome-based predictive modeling (CPM), a robust data-driven computational approach (26) to functional MRI data in order to assess associations between brain connectivity and ASD symptoms. Our primary hypothesis was that CPM would reveal a significant association between functional neuroanatomy and ASD symptoms in children with NS. Secondly, we tested the hypothesis that a model developed to predict ASD symptoms in children with and without iASD from the general population would generalize to predict ASD symptoms in children with NS. To accomplish this second goal, we included large-scale data from the Autism Brain Imaging Data Exchange (ABIDE-II). Importantly, both the NS and idiopathic cohorts were enriched for ASD symptoms. Successful cross-diagnostic prediction of ASD symptoms in the NS cohort would suggest generalizability of brain-behavior associations and similar functional brain networks underlying ASD symptoms due to RAS-MAPK upregulation and non-syndromic causes.

## Methods

### Participants

#### NS cohort

Participants with NS were recruited across the United States and Canada via caretaker-led organizations, physician referrals, and online advertisements. Eligible participants included 39 children with NS aged 4-12 years (mean=8.44, SD=2.20, 25 females). Participants were required to present results from genetic testing showing the presence of *PTPN11* (N = 29) or *SOS1* (N = 10) mutations. Complete inclusion and exclusion criteria are in the **Supplement**. This research was conducted at the Stanford University School of Medicine. The Institutional Review Board (IRB) approved all study procedures. Legal guardians completed informed consent. Participants over the age of 7 years completed assent. Children with NS were representative of the United States as illustrated in Supplemental Figure 1.

#### Non-syndromic cohort (ABIDE)

Data were gathered from the Autism Brain Imaging Data Exchange II (ABIDE) a publicly available multisite dataset of clinical, demographic, and resting-state fMRI data collected in children with ASD and typically developing (TD) (4). Participants in the ABIDE dataset were recruited from clinics and the community from multiple sites around the world (fcon_1000.projects.nitrc.org/indi/abide/). The ABIDE cohort is enriched for iASD and the present study includes data from 122 children with iASD and 230 children with TD. Local IRBs approved all study procedures and informed consent procedures were followed.

### Behavioral assessment

The Social Responsiveness Scale (SRS) was used in both NS and ABIDE to estimate social impairments and their severity, i.e., a higher score indicates more severe ASD symptoms.^27^ Raw scores were utilized to maximize individual variability and avoid differences in scoring between SRS versions (27). Full scale IQ was assessed in the NS cohort via the Wechsler Intelligence Scale for Children (WISC) (28). In the ABIDE, intelligence was assessed via the WISC, the Kaufman Brief Intelligence Test (29) or the Differential Abilities Scale (30).

### Image acquisition and preprocessing for the NS cohort

Prior to the MRI scan, all participants completed behavioral training in a mock MRI scanner to minimize motion and sensitivity to the scanner environment (31). Further imaging details are provided in the **Supplement.**

### Connectome-based predictive modeling (CPM) within each dataset

For each subject, a 268 × 268 connectivity matrix was calculated according to a previously defined functional atlas (32). The connectivity matrix was calculated using Pearson correlation coefficients of the time-courses between node pairs, followed by application of Fisher’s r-to-z transformation. Each entry in the matrix represents an ‘edge’ i.e., the strength of the functional correlation between a node pair. The complete matrix represents the individual’s functional connectome. The matrices for each subject were entered into the connectome-based predictive modeling (CPM) procedure as described in the **Supplement**.

### Application of non-syndromic ABIDE models to the NS dataset

We leverage a predictive model that was previously generated for the non-syndromic cohort (ABIDE) (33) following the same steps as described for the NS cohort. This model demonstrated a significant relationship between predicted and actual SRS Total scores (*r*=0.32, *p*<5E-10) for the population on which it was trained. Further details are described in the **Supplement**.

### Network anatomy underlying autism symptoms

The network anatomy underlying SRS scores in the NS and ABIDE datasets was compared using the hypergeometric cumulative distribution function (hygecdf, MATLAB). The probability that *n* shared edges exist between the CPM-defined network and edges within or between 10 predefined functional atlas networks (32,34) was calculated. To determine the extent to which the ABIDE and NS models share common features, the ABIDE and NS networks were compared at a ‘low threshold’ (any edge identified in any participant) and at a ‘high threshold’ (edges that appear in at least 90% of participants). The likelihood (1 – *p*-value) that each network (and between-network pair) contributed ABIDE and NS networks are reported in the Results.

## Results

Twenty-eight participants with NS were included following data scrubbing (mean framewise displacement after scrubbing = 0.147 mm, SD =0.044 mm). We include ABIDE data from 122 ASD and 230 TD after data scrubbing (33). **Table 1** shows the demographics of the NS and ABIDE cohorts.

**Table 1.**
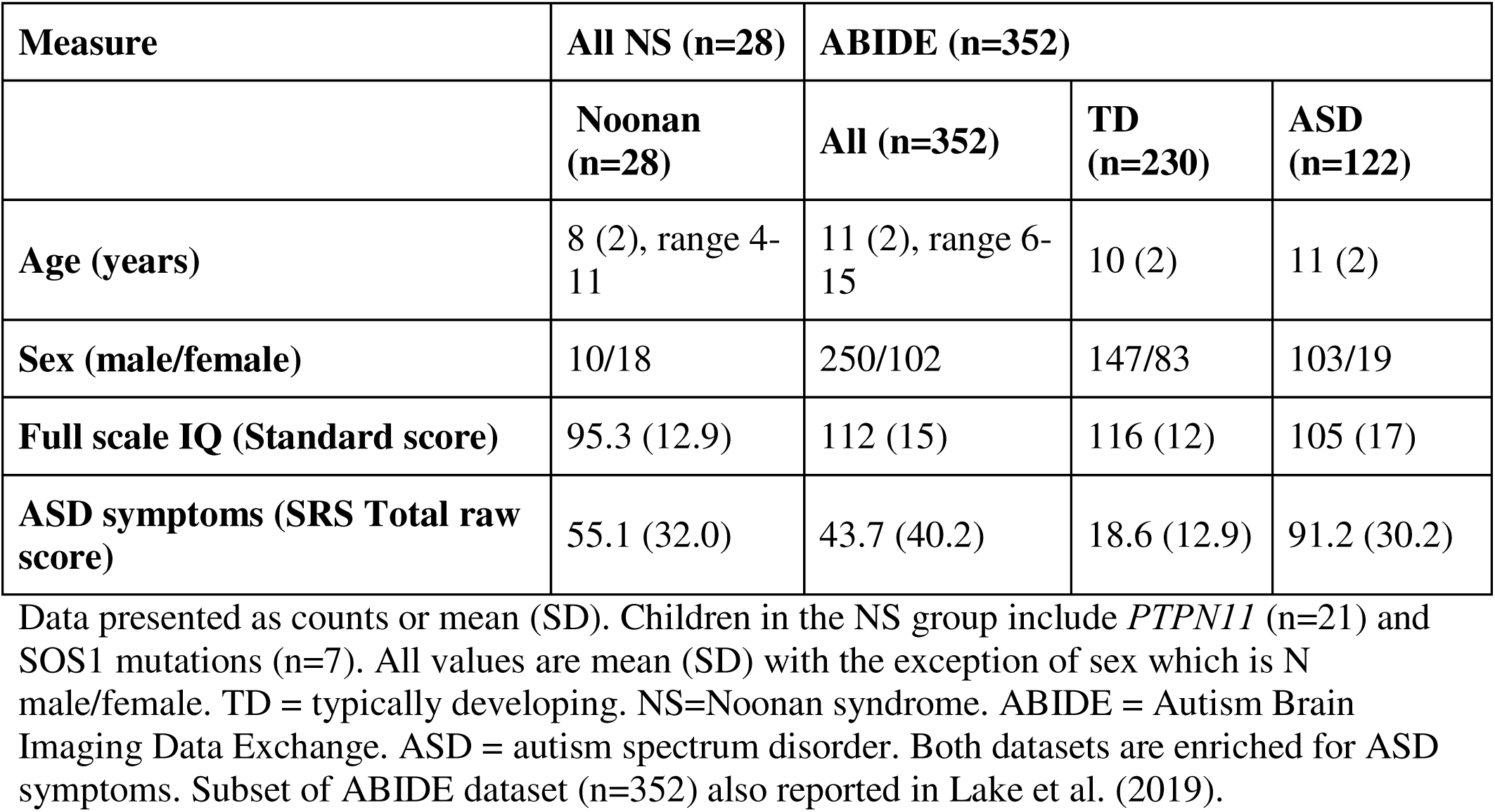
Descriptive statistics for children included in imaging analysis.

### Prediction of ASD symptoms in NS

Predicted SRS scores were significantly associated with observed SRS scores at three edge selection thresholds: *p*<.05 (*r_s_*=0.53, *p*=.041; **Figure 1)**, *p*<.01 (*r_s_*=0.43, *p*=.011), and *p*<.005 (*r_s_*=0.43, *p*=.018); *p* values were generated by comparing the correlations to the null distribution as described in the **Supplement** and thus serve as internal validation of the predictive model (**Figure 1B**.) The predicted SRS scores at each edge threshold were similar to the actual SRS scores (M=55.1, SD=32.0) but had lower variance: *p*<.05 (M=55.5, SD=14.9), *p*<.01 (M=56.2, SD=14.2), and *p*<.005 (M=55.9, SD=13.5). Supplementary analysis, including FSIQ as a covariate (alongside age and motion), indicated similar results **(Supplementary Tables 4, 5, and 6)**.

**Figure 1.**
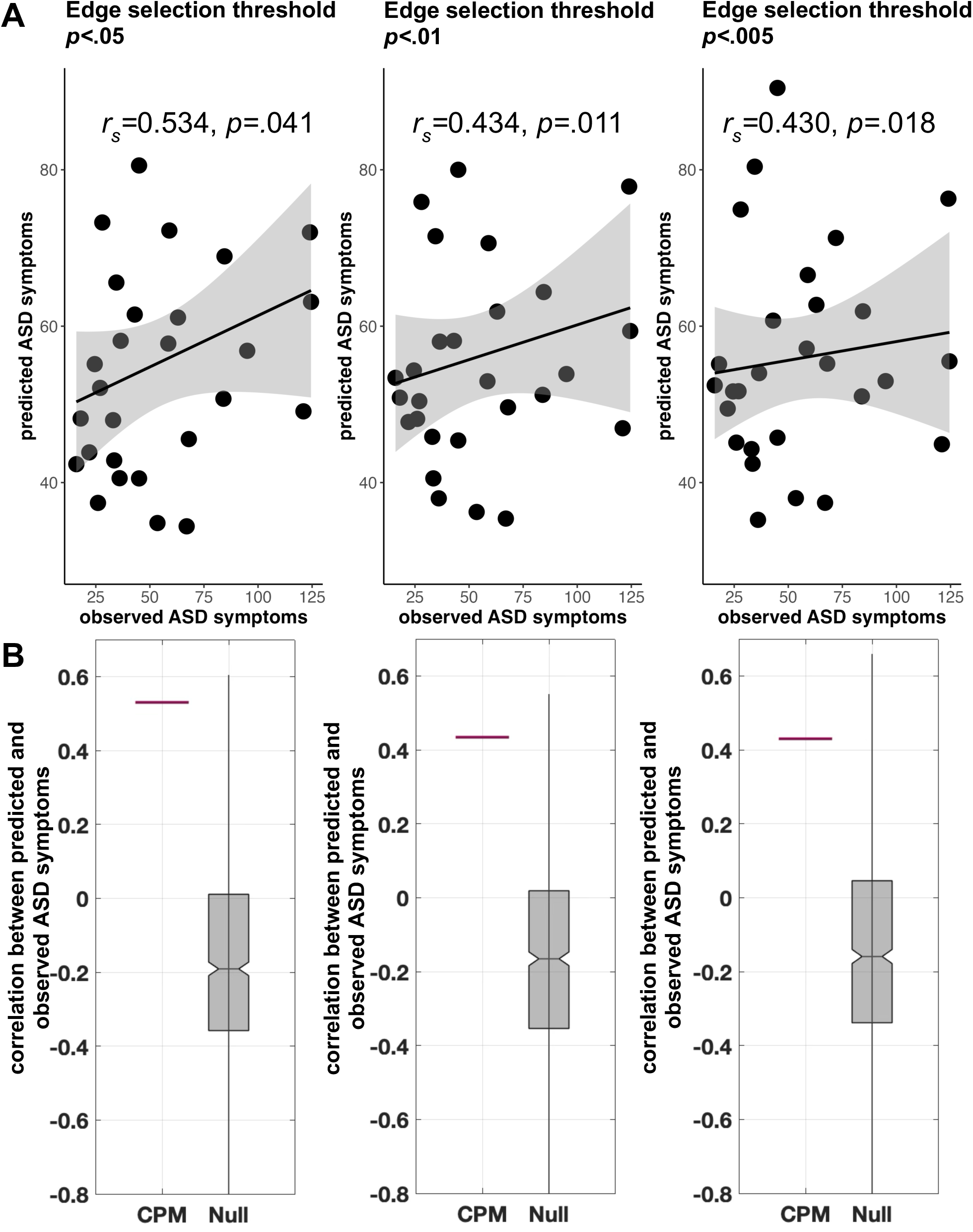
**A) Leave-one-subject-out cross-validation connectome-based predictive modeling (CPM) results in the Noonan syndrome cohort.** The sums of predicted ASD symptoms (SRS Total scores) from positive and negative models are plotted against observed ASD symptoms (SRS Total scores) at different initial edge selection thresholds of *p*<.05, *p*<.01, and *p*<.005. Spearman partial correlations (controlling for age and in-scanner motion) are presented; *p-*values indicate the significance from permutation testing (n=1000). On each plot, black line is a linear regression line and gray indicates 95% confidence interval. **B) Permutation testing indicated that correlations generated by CPM were higher than those generated by the null distribution across all edge thresholds.** In each plot, the horizontal red line shows the correlation generated by leave-one-out cross-validation CPM. The grey box shows the correlations produced when subjects and SRS Total scores are randomly shuffled prior to leave-one-out cross-validation, forming a null distribution (n=1000 permutations). ASD = autism spectrum disorder; SRS = Social Responsiveness Scale. Note that we used spearman correlation because we do not necessarily expect a linear relationship. Nonetheless, linear relationships are displayed here for visualization purposes.

### Functional anatomy of the predictive model in NS

To understand the functional anatomy of the networks, the significant edges were divided into macroscale regions. The edge threshold of *p*<.01 was used for examination of anatomy **(Figure 2**). The total number of significant positive, negative, and combined edges were 128, 214, and 342 edges, respectively. The greatest number of significant positive edges were between the subcortical-cerebellar networks (45 edges). The greatest number of significant negative edges were between visual area I-visual association area (47 edges), and the frontoparietal-motor areas (39 edges).

**Figure 2.**
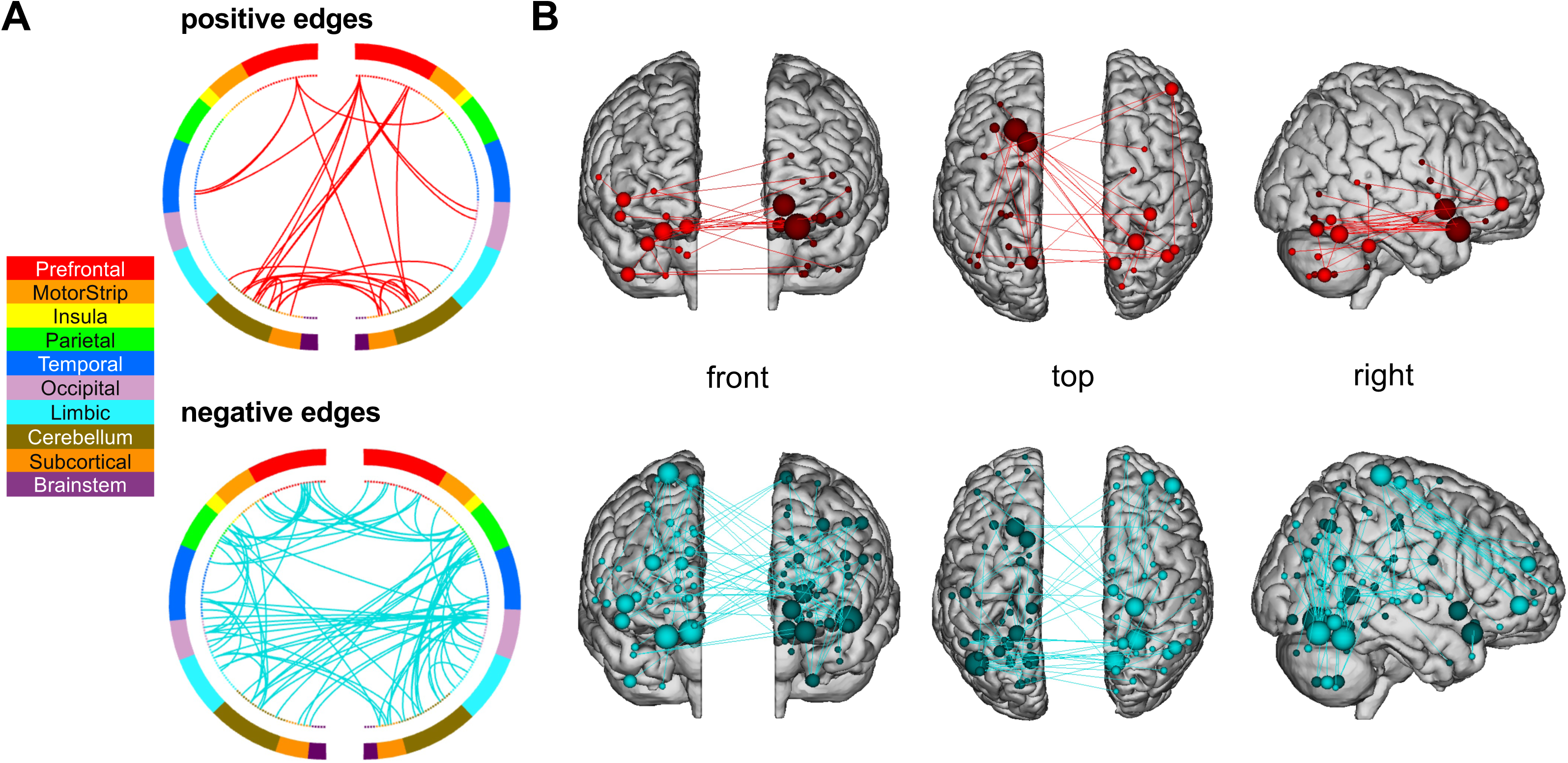
Functional networks predictive of ASD symptoms in children with NS. **A)** Connectivity patterns between networks generated by the predictive model. **B)** Anatomy of significant edges. Positive edge networks presented in red and negative edge networks presented in blue. Edges were selected at a significance threshold of *p*<.01. ASD = autism spectrum disorder; ND = Noonan syndrome. Figure created using https://bioimagesuiteweb.github.io/webapp/connviewer.html

### Cross-dataset prediction: application of ABIDE model to NS

The predictive model generated in ABIDE significantly predicted ASD symptoms in NS (*r_s_*=0.460, *p*=.018, **Figure 3**). The relationship between predicted and observed SRS Total score in the NS cohort was significant at all tested edge selection thresholds (25%, 50%, 75%, 90%, 100%); Spearman partial correlations ranged from *r_s_*=0.438 to 0.460, *p*=.025 to .018. **Figure 3** shows the correlation at an edge selection threshold of 90%, meaning only edges significant in 90% of subjects were included.

**Figure 3.**
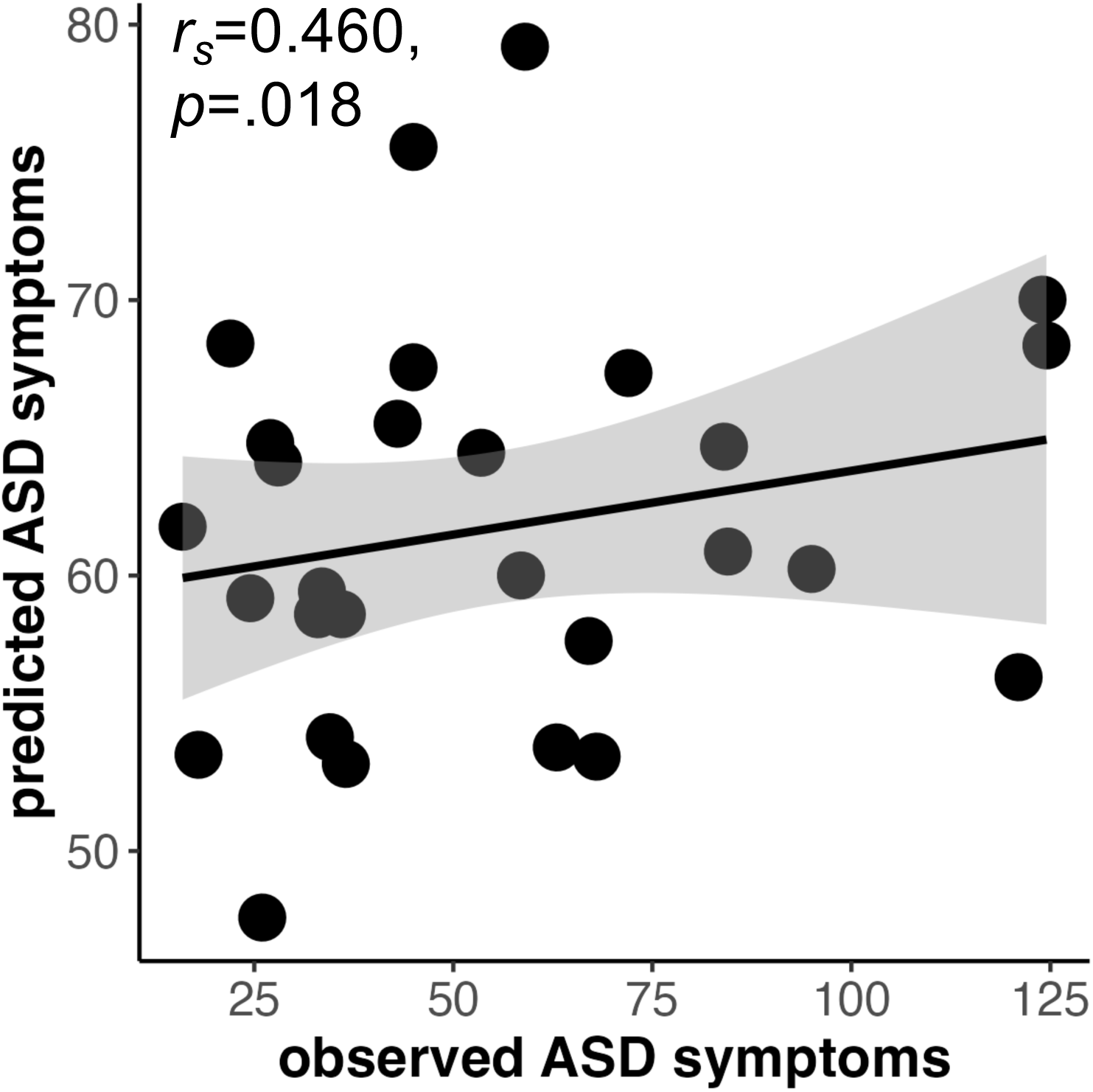
The predictive model generated using ABIDE predicts ASD symptoms in an NS cohort. A Spearman partial correlation including covariates of motion and age was used to assess the relationship between predicted and true ASD symptoms in the NS cohort following application of the predictive model from ABIDE. We used Spearman as a non-parametric correlation as we did not assume a linear relationship. Only edges significant in 90% of subjects were included in the model. ASD=autism spectrum disorder; NS=Noonan syndrome. ABIDE = Autism Brain Imaging Data Exchange.

### Consistency of model prediction between within NS models and ABIDE applied to NS models

We also compared the accuracy of model prediction when using within group NS CPM vs the ABIDE model to predict in NS. When the SRS Total score predicted by NS CPM deviates from the actual score, the SRS Total score predicted by ABIDE also deviates from the actual score (*r_s_*=0.916, p<.001; **Supplementary Figure 3**).

### Anatomy of ASD symptom networks in NS and ABIDE

Network edges were summarized according to a previously defined functional atlas(35) to capture the contribution of individual subnetworks. The likelihoods that each within-network or between-network edge contributed to ASD symptom networks are shown in **Figure 4** and **Supplementary Figure 4**. Shared and distinct network anatomy are described in **Supplemental Results.**

**Figure 4.**
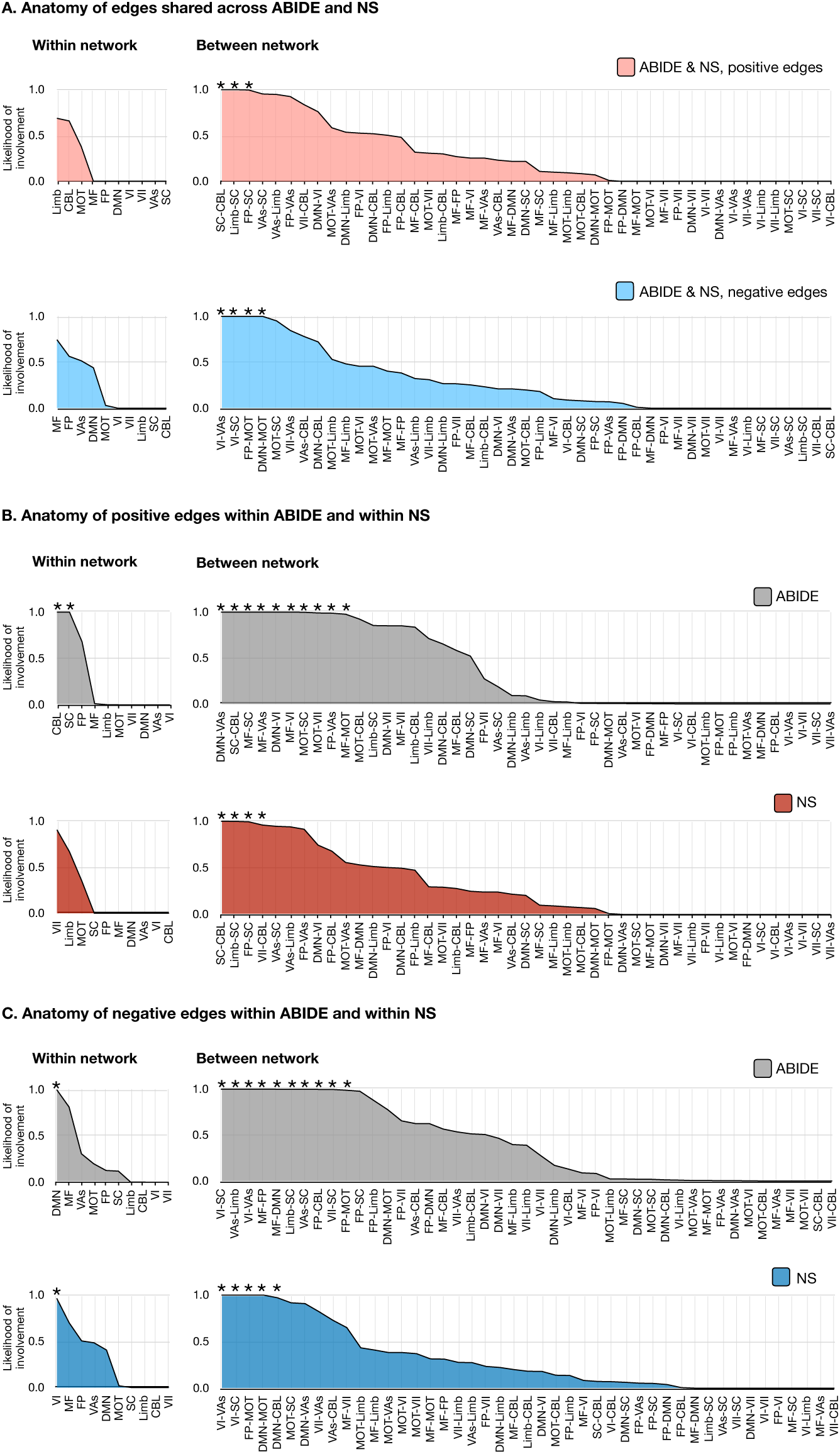
Anatomy of ASD symptom networks in ABIDE (n=352) and NS (n=28). The shared edges appearing in both ABIDE and NS are summarized in **(A)** with positive edge networks in pink and negative edge networks below in blue. The anatomy of edges within ABIDE and within NS are summarized in **(B) for positive edges,** with ABIDE shown in gray and NS shown in red and in (C) for negative edges, with ABIDE shown in gray and NS shown in blue. These plots show the likelihood (1.0 – *p*-value) of edges being shared between *a priori* networks and the ASD symptoms network for 90% of participants in each of the cohorts. In all plots, networks and between-network pairs are ordered from greatest to least cumulative likelihood. These network representations were calculated using the hypergeometric distribution function in MATLAB. * indicates a likelihood greater than chance, i.e., *p*<.05. ASD=autism spectrum disorder; NS=Noonan syndrome, ABIDE = Autism Brain Imaging Data Exchange; CBL, cerebellum; DMN, default mode network; FP, frontoparietal lobe; Limb, limbic system; MF, mediofrontal cortex; MOT, motor areas; SC, subcortical areas; VAs, visual areas; VI, visual; VII, visual-II.

## Discussion

We probed the functional architecture underlying autism symptom severity in children with NS and a non-syndromic cohort, both enriched for ASD. Functional connectivity, particularly between subcortical-cerebellar and visual regions, significantly predicted autism symptom severity in children with NS. This neural circuitry may represent an endophenotype that links the effects of RAS-MAPK pathway pathogenic variants to autism symptoms and builds upon genetic studies which previously identified a role of the RAS-MAPK pathway in idiopathic ASD (8,36). Cross dataset prediction indicated significant overlap in the brain circuitry underlying autism symptoms in children with and without NS. Using models developed in the two independent cohorts, brain network similarities between NS and the general population were limited to between-network connections; within-network connections were not shared. These findings highlight the utility of examining relatively rare genetic syndromes in the context of large-scale data from the general population to disentangle the effects of specific pathways such as RAS-MAPK on brain functioning and resultant autism symptoms. This line of work holds promise for translating neuroscience findings into clinically meaningful breakthroughs.

### Examining the effects of RAS-MAPK pathway on functional connectivity and resulting autism symptoms

We demonstrate a unique pattern of neural circuitry underlying autism symptoms in children with NS. A significant proportion of the underlying network anatomy was localized to edges connecting subcortical and cerebellar regions, indicating a positive association between functional connectivity in these regions and ASD symptoms. Subcortical regions are often highlighted in studies of NS due to structural and functional alterations. Structural MRI studies demonstrated reduced subcortical volumes in children with NS (37,38). Furthermore, advanced diffusion MRI models revealed reduced neurite density in these regions, suggesting impaired microstructural integrity, particularly in the hippocampus, putamen, and thalamus (39). Complementing these structural findings, seed-based functional connectivity analyses indicate hypoconnectivity between subcortical regions in NS (25). These results indicate convergence of structural and functional disruptions in subcortical regions in children with NS. Further research is needed to determine whether structural connectivity also contributes to ASD symptoms, which could ultimately provide a more comprehensive understanding of the neural mechanisms underlying NS and ASD more broadly.

The network anatomy underlying autism symptoms in NS revealed reduced connectivity (negative edges) between primary visual and visual association areas, as well as between frontoparietal and motor areas. Reduced connectivity in visual networks may contribute to deficits in processing and integrating visual information, aligning with known visuospatial impairments in NS (40). Similarly, altered frontoparietal-motor connectivity may underlie motor coordination, executive function, and social difficulties. Given motor impairments in NS (41), future research may explore how these connectivity alterations evolve across development and link to visuospatial and motor coordination difficulties.

### Transdiagnostic links between neural circuitry and autism symptoms

Our results reveal a common positive predictive network of neuroanatomy, including subcortical and cerebellar regions, transdiagnostically in children with and without NS. This underscores the role of the cerebellum in the ‘social brain’ across a wide range of ASD symptoms and suggests that cerebellar involvement may occur outside the involvement of the RAS-MAPK pathway. Accumulating evidence supports the involvement of the cerebellum in social functioning, and several studies have documented alterations in cerebellar structure among individuals with ASD (42–44). In contrast, research exploring the cerebellum in NS has primarily concentrated on tumors, which are highly prevalent in NS (45,46). Our group’s diffusion MRI studies in NS identified significant structural alterations in the cerebellum, including reduced fractional anisotropy (22) and decreased neurite density relative to TD peers (39). Children with ASD have demonstrated comparable reductions in fractional anisotropy (47,48) and neurite density (49), indicating shared transdiagnostic structural alterations in the cerebellum. Given these parallels, future research may explore the relationship between structural connectivity in NS and its implications for social impairment.

We also identified a common negative predictive network indicating that less connectivity was associated with greater ASD symptoms. This network included primary visual and the visual association area, primary visual and the subcortical regions, frontoparietal and motor areas, and the default mode network (DMN) and motor area. Weaker connectivity has been found previously in children and adolescents with ASD, particularly in areas of the DMN (4). Several studies have also found associations between weaker functional connectivity in the DMN and greater symptom severity (50,51), (i.e., individuals with weaker DMN connectivity received higher scores on the Autism Diagnostic Observational Scale (ADOS) and the SRS-Total) (52). Previous work in NS revealed areas of both hyper and hypoconnectivity (23,25); however, these studies did not investigate associations with autism symptoms. Our findings suggest similar patterns of predictive between-network connectivity in children with NS and ASD, suggesting non syndromic samples may be leveraged to better understand the functional neuroanatomy underlying autism symptoms in NS.

A major distinction between NS and non-syndromic cohorts was the lack of convergence for within-network connectivity. Only between-network connectivity patterns were shared across cohorts. This finding speaks to the importance of examining connectivity patterns outside the boundaries of traditionally defined resting state networks, which is not always the norm in functional neuroimaging studies, including our recent work in NS (25). In fact, within the NS cohort, the pattern of connectivity underlying autism symptoms was primarily between network connections. We previously found globally increased within-network connectivity for children with NS relative to children with TD and interpreted this as compensation for less efficient connectivity (25). A predominance of between network and less within network connectivity, as found in the present study, may indicate additional reorganization due to inefficient network processing. Taken as a whole the divergence between connectivity patterns among NS and non-syndromic cohorts indicates the breadth of distinct neurobiological processes that may underlie autism symptoms in each condition.

## Limitations

Image protocols and preprocessing were tailored to the unique aspects of each dataset and used rigorous data censoring, yet there were differences between the two cohorts. A recent preprint suggests resting-state fMRI pipeline variation has limited effects on data outcomes (53) although we cannot rule out unintended effects. The age ranges of the NS (4 to 11 years) and non-syndromic (6 to 15 years) cohorts were different; however, age was included as a covariate in all analyses. Future work with more accurately matched age cohorts and longitudinal follow-up will be important to understand how networks underlying ASD symptoms change with developmental processes. The size of our NS cohort is small, given the relative rarity of the NS mutations. Thus, we used a leave-one-out cross-validation and compared the result to a permutation-based null distribution. For a larger sample size, a split-half train-test split would be a more rigorous approach. However, the significant cross-dataset comparison yielded in this study suggests that use of large publicly available datasets (such as ABIDE) can be useful in contextualizing smaller and harder to collect datasets in rare genetic syndromes.

## Conclusion

Leveraging data-driven methodology, we uncovered similarities and distinctions in the predictive brain networks associated with ASD symptoms among children with and without NS. The presence of shared brain networks suggests a converging pattern of functional connectivity underlying ASD symptoms in both NS and iASD. According to recent genetic work, this pattern may signify the common involvement of the RAS-MAPK signaling pathway in ASD more broadly and points to treatments aimed at targeting RAS-MAPK as potentially promising therapeutic avenues.

## Data Availability

All data produced in the present study are available upon reasonable request to the authors.

https://fcon_1000.projects.nitrc.org/indi/abide/

## Abbreviations

ABIDE: Autism Brain Imaging Data Exchange
ASD: autism spectrum disorder
CPM: connectome-based predictive modeling
DMN: default mode network
fMRI: functional magnetic resonance imaging
iASD: idiopathic autism spectrum disorder
NS: Noonan syndrome
RAS-MAPK: rat-sarcoma mitogen-activated protein kinase
SRS: Social Responsiveness Scale
TD: typical developing
WISC: Wechsler Intelligence Scale for Children

## Declarations

### Ethics approval and consent to participate

This research was conducted at the Stanford University School of Medicine. The Institutional Review Board (IRB) approved all study procedures. Legal guardians completed informed consent. Participants over the age of 7 years completed assent.

### Consent for publication

Only aggregate data is presented. Data from an individual person are not presented.

### Availability of data and materials

The final dataset collected for this manuscript will be stripped of identifiers and made available to qualified researchers. Data from the Autism Brain Imaging Data Exchange is available here: https://fcon_1000.projects.nitrc.org/indi/abide/

### Authors’ contributions: Jennifer Bruno

Conceptualization; Formal Analysis; Funding Acquisition; Investigation; Software; Writing – Original Draft Preparation. **Julia Plank:** Formal Analysis; Software; Visualization; Writing – Original Draft Preparation. **Sam Leder:** Writing – Original Draft Preparation. **Evelyn Lake:** Conceptualization; Resources; Data Curation; Methodology; Resources; Software. **Emily Finn:** Conceptualization; Resources; Methodology; Resources; Software. **Tamar Green:** Conceptualization; Funding Acquisition; Supervision. All authors contributed to review and editing of the manuscript.

## Acknowledgements

We thank the families who participated in this research. The authors would also like to thank the Noonan Syndrome Foundation, the RASopathies Network, and the Children’s Tumor Foundation which made this work possible. We would like to thank Stanford University and the Stanford Research Computing Center for providing computational resources and support that contributed to these research results, some of the computing for this project was performed on the Sherlock cluster. We gratefully acknowledge the support of The Lucas Service Center at Stanford.

