## Supplement for "Transdiagnostic similarities and distinctions in brain networks associated with ASD symptoms: A prospective cohort study"

**Supplementary Material**

**Supplemental Methods**

### ABIDE I/II

ABIDE I/II are publicly available multi-site datasets including resting-state fMRI, clinical, and demographic data. The details for the ABIDE I/II datasets are available at https://fcon_1000.projects.nitrc.org/indi/abide/abide_I.html and https://fcon_1000.projects.nitrc.org/indi/abide/abide_II.html.

### Noonan Syndrome

#### *Participant recruitment and exclusion criteria*

Participants in the present paper overlap with those presented in a previous paper which examined associations between seed-based connectivity and cognitive phenotypes.[1] Unlike prior work, the current study aims to identify functional brain networks predictive of ASD symptoms and assess the cross-dataset application of a predictive model generated in the general population to children with NS. Participants with NS were recruited via the National Noonan Syndrome Foundation, a local network of physicians and advertisements posted on the Stanford University School of Medicine website. TD participants were recruited through two NIH funded studies (HD049653 and MH099630)[2] using local print media and parent networks. Potential participants across both groups were excluded for premature birth (gestational age <32 weeks), low birth weight (<2000g) or diagnosis of a major psychiatric disorder. Additional exclusion criteria included history of the following: seizures, neurological disorders known to have an impact on cognitive development or brain structure, gross structural brain malformations. All participants were free from MRI contraindications.

#### *Image Acquisition*

Imaging data were acquired on a GE Healthcare Discovery 3.0 Tesla whole-body MR system using a standard 8-channel head coil (GE Medical Systems, Wilwaukee, WI). Whole-brain resting-state fMRI data were acquired using a T2-weighted gradient-echo spiral sequence and high-order shimming with an acquisition time (TA) of 6 minutes 8 seconds. Participants were instructed to relax and remain still. High-resolution T1-weighted structural images were acquired using a magnetization-prepared rapid gradient-echo (MPRAGE) sequence.

#### *Image Preprocessing*

Image preprocessing was conducted using *fMRIPrep* 1.3.0.post3 (RRID:SCR_016216) . To minimize the impact of motion artifacts, frames that were displaced by > 0.5 mm were eliminated along with the prior frame and the two subsequent frames.[3] To be included in analysis, imaging data was also required to meet the following two conditions: 1) the quality and orientation of structural and functional scans were deemed sufficient as outlined in registration procedures published by FMRIPrep https://fmriprep.org/en/1.0.3/api/index.html; and 2) a minimum of 4 minutes of functional data was artifact-free following frame elimination as described above. Following this procedure, twenty-eight of the 39 eligible participants were included in further analysis.

For each participant, a reference functional volume and its skull-stripped version were generated using *fMRIPrep*. A deformation field was estimated based on *fMRIPrep*'s *fieldmap-less* approach to correct for susceptibility distortions. The deformation field was generated by co-registering the BOLD reference to each participant's T1w-reference with its intensity inverted.[4] Registration was performed with antsRegistration (ANTs 2.2.0), and the process was regularized by constraining deformation to be nonzero only along the phase-encoding direction, and modulated with an average fieldmap template.[5] Based on the estimated susceptibility distortion, an unwarped BOLD reference was calculated for a more accurate co-registration with each anatomical reference. The BOLD reference was then co-registered to each T1w reference using flirt (FSL 5.0.9)[6] and the boundary-based registration cost-function.[7] Co-registration was configured with nine degrees of freedom to account for distortions remaining in the BOLD reference. Head-motion parameters with respect to the BOLD reference (transformation matrices, and six corresponding rotation and translation parameters) were estimated before spatiotemporal filtering using mcflirt (FSL 5.0.9).[8] BOLD images were slice-time corrected using 3dTshift (AFNI 20160207 RRID:SCR_005927).[9] The preprocessed BOLD time-series were resampled onto their original, native space by applying a single, composite transform to correct for head-motion and susceptibility distortions. The BOLD time-series were resampled to MNI152NLin2009cAsym standard space, generating a *preprocessed BOLD run in MNI152Nlin2009cAsym space*. Several confounding time-series were calculated based on the *preprocessed BOLD*: framewise displacement (FD), DVARS (D is the temporal derivative of the time course, and VARS refers to root mean square variance over voxels) and three region-wise global signals. FD and DVARS are calculated for each functional run, using *Nipype*.[3,10] The three global signals are extracted within the CSF, the WM, and the whole-brain masks. Gridded (volumetric) resamplings were performed using antsApplyTransforms (ANTs), configured with Lanczos interpolation to minimize the smoothing effects of other kernels.[11] Non-gridded (surface) resamplings were performed using mri_vol2surf (FreeSurfer). Finally, a temporal band-pass filter (0.01 Hz < f < 0.1 Hz) was applied.

**Image acquisition and preprocessing for the non-syndromic ABIDE cohort**
All acquisition and analysis steps for this cohort are detailed here.[12] All scanning was completed on a GE Healthcare Discovery 3.0 Tesla whole-body MR system using a standard 48-channel head coil (GE Medical Systems, Wilwaukee, WI). Whole-brain functional images were collected using a T2-weighted gradient-echo spiral sequence and high-order shimming (echo time=30 ms; repetition time=2,000 ms; flip angle=80°; field of view=22 cm x 22 cm; acquisition matrix=64x64; approximate voxel size=3.4x3.4x5mm). Participants were instructed to relax and remain still in the scanner with their eyes closed during the 6 min 8 second resting state scan. Four frames were automatically discarded at scanner and 5 frames were subsequently removed during preprocessing; thus 175 frames were available for subsequent data analysis.

**CPM procedure details**
CPM was applied to the NS cohort using the following procedures. High-resolution T1-weighted (T1w) structural images were also collected during the same session to facilitate normalization to standard space (sagittal slices, repetition time 8.2 ms; echo time 3.2 ms; flip angle 12°; field of view 256 × 256 mm; matrix 256 × 256; 176 slices; voxel size = 1.0 × 1.0 × 1.0 mm).

CPM was used to assess the relationship between brain connectivity and SRS scores.[13,14] Briefly, CPM is a data-driven predictive modeling approach that aims to identify patterns of functional brain connectivity that predict behavior (here, ASD symptoms quantified by the SRS Total score) in novel subjects. A leave-one-out cross-validation was used to build the predictive model. This process involves, on each iteration, removing one subject from the dataset, training a model on the remaining subjects (training data), then using this model to predict the score for the left-out (test) subject. The steps involved are summarized as follows. 1) Individual 268 × 268 connectivity matrices and SRS Total scores were loaded into MATLAB version R2024. 2) To determine the edges most significantly related to SRS score, Spearman partial correlations between each edge and SRS score were performed on the training data. In all models within the NS data, age and in-scanner motion (expressed as average framewise displacement in mm) were included as covariates. For all models within the population-based cohort, age, in-scanner motion and full scale IQ standard score were included as covariates. Full scale IQ was not used as a covariate in the primary analysis within the NS group to avoid overfitting given the small sample size. Subsequent analysis was performed with full scale IQ covariate for verification that this difference did not drive any of the between group differences. The initial edge selection threshold was varied ($p$<.05, .01, .005 associated with the partial Spearman correlation) to examine the effect on model accuracy. 3) Positive and negative networks were constructed. Positive networks included edges whose connectivity was positively associated with SRS scores. Negative networks included edges whose connectivity was negatively associated with SRS scores. 4) For each subject in the training data, the edge weights are summed within each network (positive and negative) to produce a single subject value for each network. 5) A linear model was fit to the training data separately for the positive and negative networks. Single-subject summary values were calculated for the test subject separately for the positive and negative networks, and these values were used as input to the predictive model to generate a predicted SRS score for the test subject. 6) Following all iterations (i.e., once all subjects had been held out once), model accuracy was computed using the Spearman correlation between predicted and observed SRS scores. 7) The accuracy observed in step (6) was evaluated for statistical significance using permutation testing ($n$=1000 permutations). The SRS scores were randomly shuffled across subjects to break the true brain-behavior relationship and a null distribution was generated by predicting SRS scores (i.e., following steps 1-6) based on the shuffled data. $P$-values of the permutation tests were calculated based on the proportion of sampled permutations that were greater or equal to the true prediction correlation. Networks were visualized using the open-source BioImage Suite Web software (https://bioimagesuiteweb.github.io/webapp/) recommended in the CPM protocol.[13]

**Application of non syndromic ABIDE models to the NS dataset**

The parameters from this previously generated model were applied to the NS cohort using the following steps. First the 268 × 268 matrices and SRS Total scores from the NS subjects were loaded into MATLAB. Next the results from CPM of the ABIDE dataset were loaded; these results included the significant edges in each of the 268 × 268 matrices for each model (n=352, since there were 352 subjects in the ABIDE dataset, and models were trained via leave-one-subject-out cross validation) in each network (positive and negative). Next a threshold was applied to select the set of edges that were significantly associated with SRS scores in the ABIDE network such that only edges appearing in a certain % of participants would be retained for further analysis. Various edge selection thresholds were tested: 0.25, 0.50, 0.75, 0.90, and

1.00 (i.e., an edge threshold of 0.75 indicates only edges appearing in 75% of ABIDE subjects would be used in the analysis). Finally, the predictive model, defined as a set of edges, was applied to the NS data and the set of edges were summed across all NS participants separately for positive and negative networks. The predicted SRS scores (generated by the ABIDE model applied to NS) were compared to the actual SRS scores using a Spearman partial correlation including covariates of age and motion.

**Supplemental Results**

**Shared and distinct anatomy of ASD symptom networks in NS and ABIDE**
Across thresholds, NS and ABIDE networks share positive subcortical-cerebellum and limbic-subcortical edges.  Negative NS and ABIDE networks share between-network edges linking visual area I-visual association area, visual area I-subcortical, frontoparietal area-motor, and the default mode network-motor areas. There were no within-network edges shared between ABIDE and NS. Circuits which distinguish ABIDE from NS positive networks include edges which link subcortical-cerebellum, limbic-subcortical, default mode network-visual area I, and the mediofrontal area-visual area I. Circuits which distinguish ABIDE from NS negative networks include edges which link visual area I-subcortical, visual area I-visual association area, and frontoparietal-motor areas

| Household income | Frequency | Percent |
|---|---|---|
| </=50 | 5 | 17.9 |
| 51-100 | 9 | 32.1 |
| 101-150 | 7 | 25 |
| >/=151 | 7 | 25 |

**Supplemental Table 1.** Household income levels for each family included in the study. Income is in thousands of dollars.

**CPM prediction of ASD behavior in Noonan syndrome**

**Supplementary Table 2.** Correlations between true and predicted SRS scores derived from **positive edge networks** at different thresholds.

| ***r* threshold** | ***p* threshold** | ***r*** | ***p*** | **number of edges** |
|---|---|---|---|---|
| 0.100 | .610 | 0.268 | .045 | 12322 |
| 0.130 | .510 | 0.274 | .040 | 10214 |
| 0.150 | .450 | 0.296 | .031 | 8928 |
| 0.200 | .310 | 0.341 | .026 | 6040 |
| 0.250 | .200 | 0.351 | .029 | 3874 |
| 0.300 | .120 | 0.352 | .022 | 2352 |
| 0.320 | **.050** | **0.225** | **.079** | 730 |
| 0.480 | **.010** | **0.084** | **.226** | 128 |
| 0.520 | **.005** | **0.131** | **.149** | 60 |

**Supplementary Table 3.** Correlations between true and predicted SRS scores derived from **negative edge networks** at different thresholds.

| ***r* threshold** | ***p* threshold** | ***r*** | ***p*** | **number of edges** |
|---|---|---|---|---|
| 0.100 | .610 | 0.428 | .009 | 14418 |
| 0.130 | .510 | 0.437 | 0.010 | 11978 |
| 0.150 | .450 | 0.443 | .007 | 10552 |
| 0.200 | .310 | 0.467 | .006 | 7472 |
| 0.250 | .200 | 0.460 | .008 | 4924 |
| 0.300 | .120 | 0.490 | .005 | 3178 |
| 0.320 | **.050** | **0.530** | **.003** | 1148 |
| 0.480 | **.010** | **0.424** | **.025** | 214 |
| 0.520 | **.005** | **0.410** | **.015** | 96 |

**Supplementary Table 4.** Correlations between true and predicted SRS scores derived from **combined edge networks** at different thresholds.

| ***r* threshold** | ***p* threshold** | ***r*** | ***p*** | **number of edges** |
|---|---|---|---|---|
| 0.100 | .610 | 0.385 | .014 | 26740 |
| 0.130 | .510 | 0.414 | .012 | 22192 |
| 0.150 | .450 | 0.401 | .010 | 19480 |
| 0.200 | .310 | 0.408 | .010 | 13512 |
| 0.250 | .200 | 0.423 | .015 | 8798 |
| 0.300 | .120 | 0.479 | .008 | 5530 |
| 0.320 | **.050** | **0.534** | **.004** | 1878 |
| 0.480 | **.010** | **0.434** | **.016** | 342 |
| 0.520 | **.005** | **0.430** | **.012** | 156 |

**Supplementary Table 5.** Spearman correlations between true and predicted SRS scores derived from **positive edge networks** at different thresholds **including FSIQ** as a covariate (along with age and motion).

| ***r* threshold** | ***p* threshold** | **$r_s$** | ***p*** | **number of edges** |
|---|---|---|---|---|
| 0.320 | .050 | 0.174 | 0.118 | 734 |
| 0.480 | .010 | -0.003 | 0.301 | 124 |
| 0.520 | .005 | -0.036 | 0.426 | 56 |

**Supplementary Table 6.** Correlations between true and predicted SRS scores derived from **negative edge networks** at different thresholds **including FSIQ** as a covariate (along with age and motion).

| ***r* threshold** | ***p* threshold** | **$r_s$** | ***p*** | **number of edges** |
|---|---|---|---|---|
| 0.320 | .050 | 0.488 | 0.007 | 1132 |
| 0.480 | .010 | 0.414 | 0.012 | 210 |
| 0.520 | .005 | 0.359 | 0.041 | 110 |

**Supplementary Table 7.** Correlations between true and predicted SRS scores derived from **combined edge networks** at different thresholds **including FSIQ** as a covariate (along with age and motion).

| ***r* threshold** | ***p* threshold** | $r_s$ | ***p*** | **number of edges** |
|---|---|---|---|---|
| 0.320 | .050 | 0.581 | <.001 | 1866 |
| 0.480 | .010 | 0.451 | 0.007 | 334 |
| 0.520 | .005 | 0.375 | 0.025 | 166 |

**Model generalizability: application of ABIDE model to NS**

**Supplementary Table 8.** Correlations between true and predicted ASD symptoms in children with NS, following application of ABIDE model.

| **Edge threshold** | **Network of edges** | ***r*** | ***p*** | **number of edges** |
|---|---|---|---|---|
| 1.00 | Positive | 0.420 | .033 | 964 |
| | Negative | 0.275 | .175 | 736 |
| | Combined | 0.438 | .025 | 1700 |
| 0.90 | Positive | 0.368 | .064 | 1416 |
| | Negative | 0.362 | .069 | 1120 |
| | Combined | 0.460 | .018 | 2536 |
| 0.75 | Positive | 0.378 | .057 | 1494 |
| | Negative | 0.382 | .054 | 1194 |
| | Combined | 0.443 | .023 | 2688 |
| 0.50 | Positive | 0.383 | .054 | 1538 |
| | Negative | 0.388 | .050 | 1260 |
| | Combined | 0.449 | .022 | 2798 |
| 0.25 | Positive | 0.403 | .041 | 1612 |
| | Negative | 0.393 | .047 | 1312 |
| | Combined | 0.337 | .022 | 2924 |

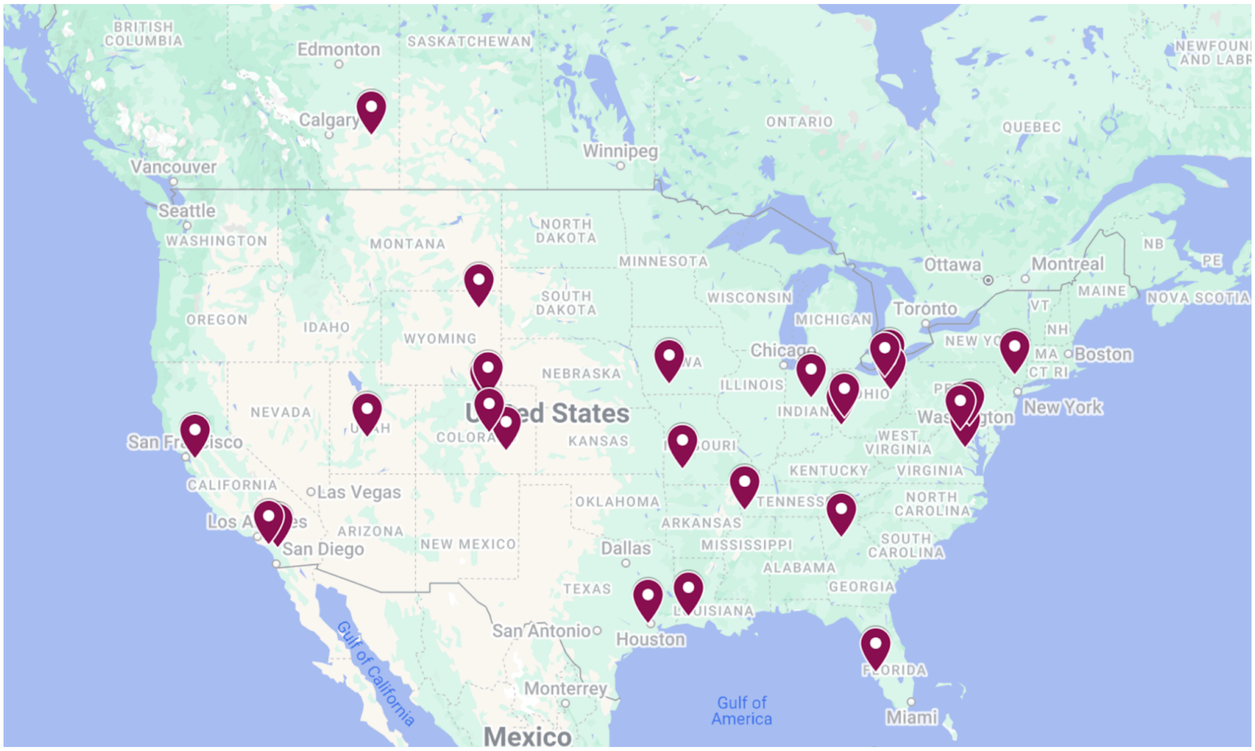


**Supplemental Figure 1.** Geographic representation of the Noonan syndrome sample demonstrates inclusion of participant families across the United States and Canada.

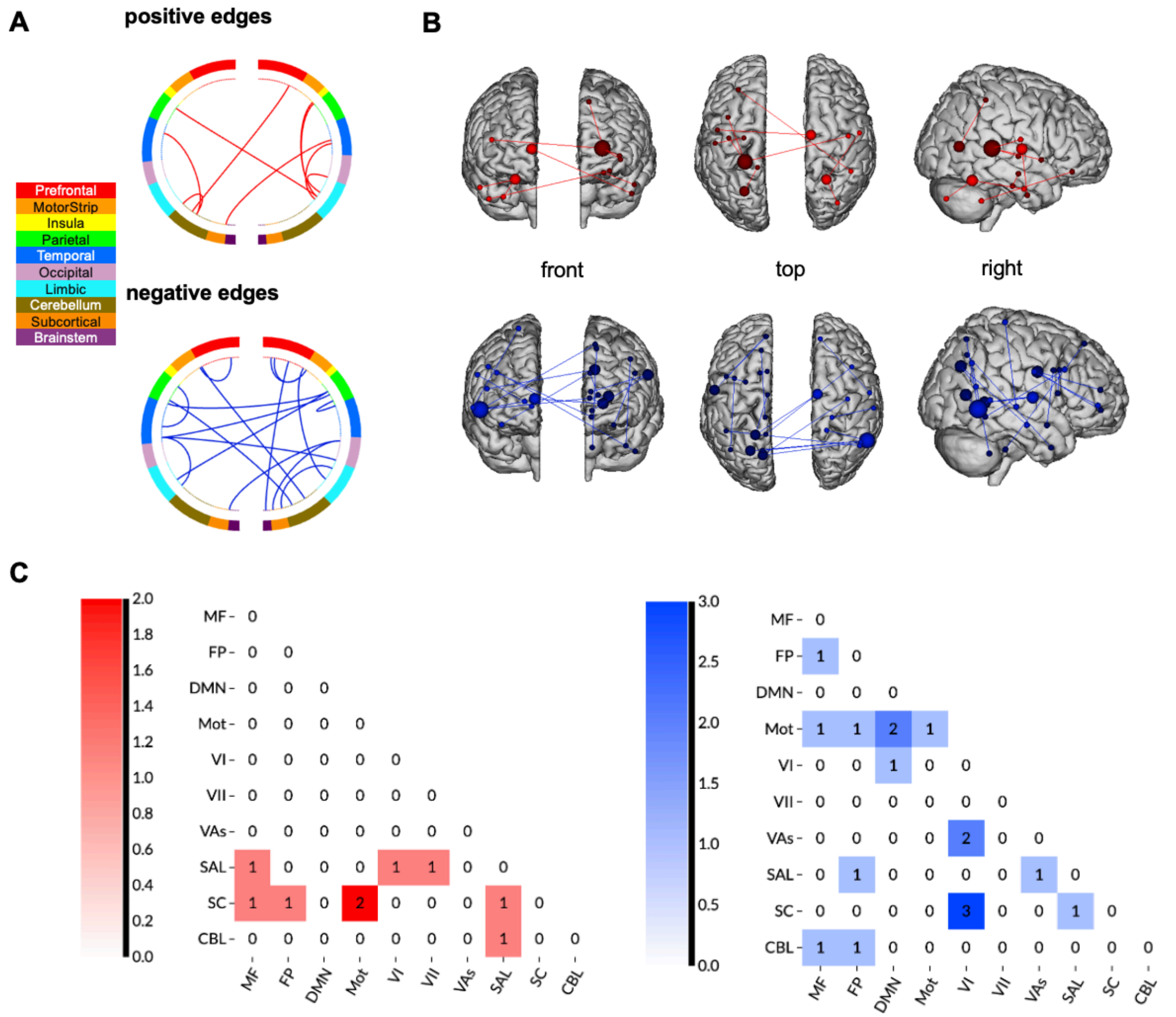


**Supplementary Figure 2.** Functional networks predictive of ASD symptoms shared between NS and ABIDE cohorts at a 25% threshold (i.e., edges significant in at least 25% of NS subjects and 25% of ABIDE subjects were compared). A) Connectivity patterns between networks generated by the predictive model. B) Anatomy of significant shared edges. C) Number of significant shared edges between and within networks. Positive edge networks presented in red and negative edge networks presented in blue. Figure created using https://bioimagesuiteweb.github.io/webapp/connviewer.html *MF=medial frontal, FP=frontoparietal, DMN=default mode network, Mot=motor, VI=visual area I, VII=visual area II, VAs=visual association, SAL=salience, SC=subcortical, CBL=cerebellum.*

In addition to comparing ABIDE and NS at the network level, we also investigated the number of overlapping edges between the groups. We found few shared edges between the groups. At a threshold of 100% (i.e., edges must overlap in 100% of NS and 100% of ABIDE), we found only 2 shared edges. At a more liberal threshold of 25% (i.e., edges must overlap in minimum 25% of NS and 25% of ABIDE), we found 26 shared edges (**Supplementary Figure 2**).

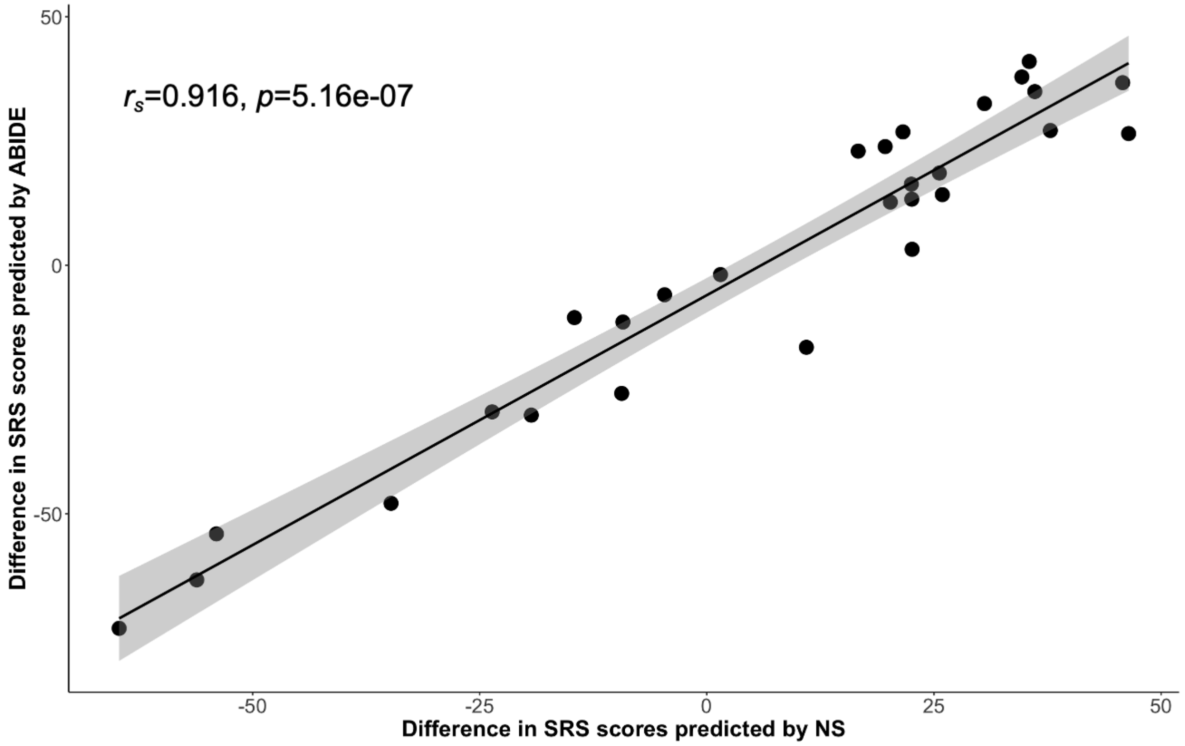


**Supplementary Figure 3.** Difference in SRS Total scores (predicted minus actual SRS Total scores for NS subjects) predicted by CPM within the NS cohort (x-axis) and predicted using the modes generated from CPM in ABIDE and applied to NS (y-axis). The Spearman correlation indicates a positive association, suggesting that when the SRS Total score predicted using models developed within the NS cohort deviates from the actual score, the SRS Total score predicted by ABIDE also deviates from the actual score.

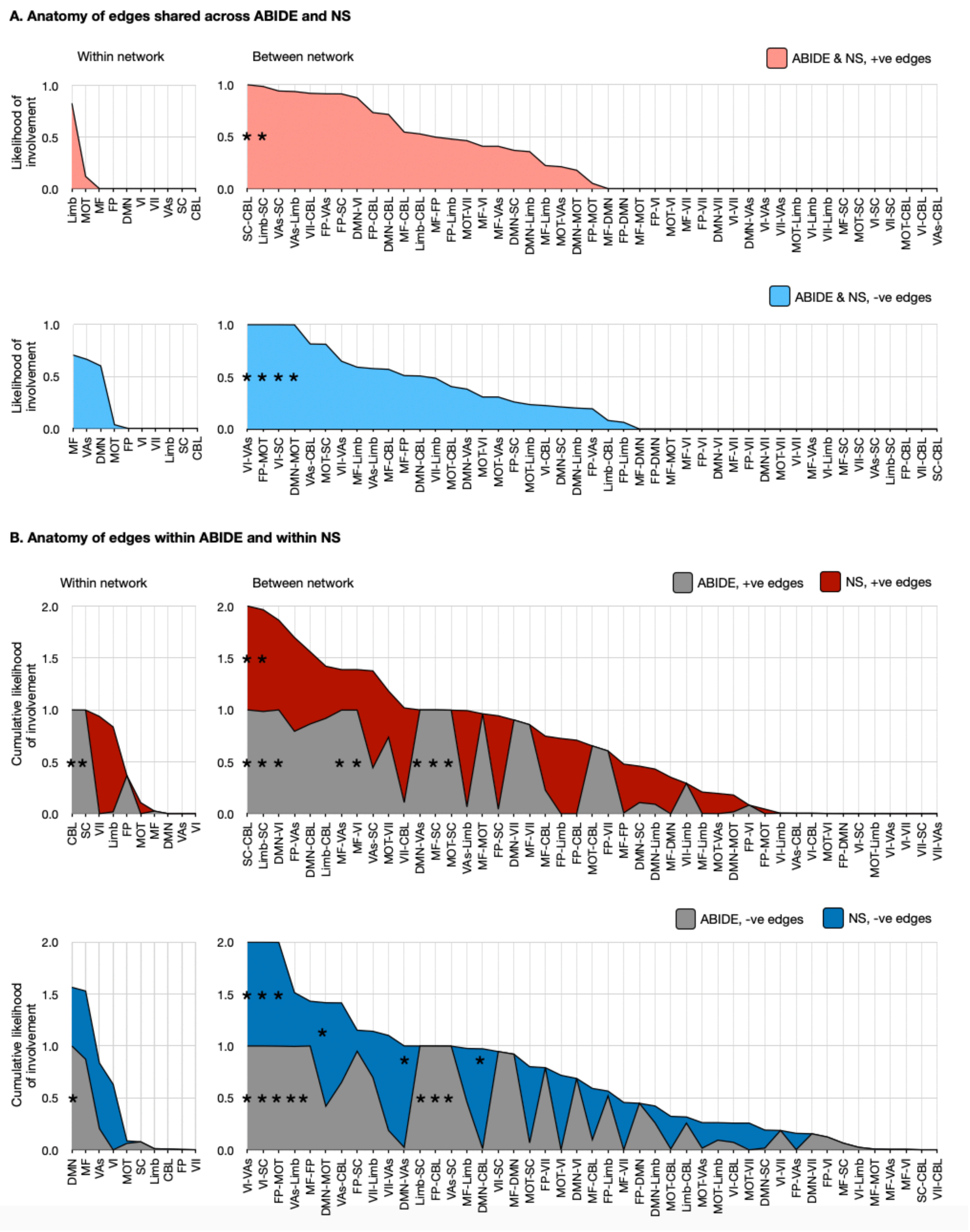


**Supplementary Figure 4.** Anatomy of ASD symptom networks in ABIDE (n=352) and NS (n=28). The shared edges across ABIDE and NS are summarized in **(A)** with positive (+ve) edge networks in pink and negative (-ve) edge networks below in blue. The anatomy of edges within ABIDE and within NS are summarized in **(B)** with ABIDE shown in gray and NS shown in color. These network representations were calculated using the hypergeometric distribution function in MATLAB. * indicates a likelihood greater than chance. -ve, negative; +ve, positive; ASD, autism spectrum disorder; CBL, cerebellum; DMN, default mode network; FP, frontoparietal lobe; Limb, limbic system; MF, mediofrontal cortex; MOT, motor areas; SC, subcortical areas; VAs, visual areas; VI, visual; VII, visual-II.
